# The Contribution of Commonly Diagnosed Genetic Disorders to Small for Gestational Age Birth and Subsequent Morbidity and Mortality in Preterm Infants

**DOI:** 10.1101/2023.07.14.23292682

**Authors:** Miles Bomback, Selin Everett, Alex Lyford, Rakesh Sahni, Faith Kim, Caitlin Baptiste, Joshua E. Motelow, Veeral Tolia, Reese Clark, Thomas Hays

## Abstract

**Objective:** Preterm infants born small, vs. appropriate for gestational age (SGA, AGA) are at greater risk for morbidity and mortality. The contribution of genetic disorders to preterm SGA birth, morbidity, and mortality is unknown. We sought to determine the association between genetic disorders and preterm SGA birth, and the association between genetic disorders and morbidity or mortality within preterm SGA infants. We hypothesized that genetic disorders were significantly associated with both.

**Study Design:** This was a retrospective multicenter cohort study of 409 339 infants, born 23–33 weeks’ gestation between 2000 and 2020. The odds of preterm SGA (vs AGA) birth, and the odds of severe morbidity or mortality within SGA preterm infants were determined for infants with genetic disorders, after adjusting for known risk factors.

**Results:** Genetic disorders were present in 3.0 and 1.3% of SGA and AGA preterm infants respectively; genetic disorders conferred an aOR (95% CI) of 2.06 (1.92, 2.21) of SGA birth. Genetic disorders were present in 4.3 of preterm SGA infants with morbidity or mortality and 2.1% of preterm SGA infants that did not experience morbidity or mortality. Genetic disorders conferred an aOR (95% CI) of 2.12 (2.66, 3.08) of morbidity or mortality.

**Conclusions:** Genetic disorders are strongly associated with preterm SGA birth, morbidity, and mortality. Clinicians should consider genetic testing of preterm SGA infants, particularly in the setting of other comorbidities or anomalies. Prospective, genomic research is needed to clarify the contribution of genetic disorders to disease in this population.

## Introduction

Preterm infants experience a profound burden of morbidity and mortality, and small for gestational age (SGA) birth is one of the strongest clinical risk factors for poor outcomes. ^1–9^ SGA is defined as birth weight less than the 10^th^ percentile or two standard deviations below mean for gestational age and sex based on estimates of population distributions.^10–13^ The mechanism(s) by which SGA birth confers clinical risk are largely unknown. Known factors including impaired metabolic reserve explain only a fraction of these outcomes.^14^ Although there are known risks for SGA birth, including maternal, placental, infectious, genetic, and environmental factors, the etiology of SGA birth is poorly understood. Known risks include maternal.^14–16^

The overall contribution of genetic disorders to preterm SGA birth is unknown. Fetal growth restriction (FGR) and continued poor fetal growth often impacts the decision to prematurely deliver a fetus. Multiple studies have found genetic disorders associated with FGR.^17,18^ Higher rates of aneuploidy and copy number variants are found in FGR, particularly when presenting before 32 weeks’ gestation or accompanied by other fetal anomalies.^19,20^ Metanalysis of exome and genome sequencing for FGR demonstrated single gene disorders in 4% of isolated FGR and 30% of FGR accompanied by other anomalies.^17^ Genetic disorders have been shown to contribute to morbidity and severe mortality in related populations as well. Genetic disorders have been found in 6% of stillbirths, 10% of fetal structural anomalies, and in 20% of cohorts of pediatric critical illness or perinatal disease.^21–26^

Given the burden of illness experienced by preterm SGA infants, emerging evidence that genetic disorders cause FGR and SGA, and evidence that genetic disorders cause critical illness in related populations, we sought to determine the contribution of commonly diagnosed genetic disorders to preterm SGA birth, morbidity, and mortality. The Pediatrix Clinical Datawarehouse (CDW) contains detailed health records of these infants, including the presence of genetic disorders diagnosed during standard clinical care.

## Materials/Subjects and Methods

This study was approved by the institutional review board of Columbia University with a waiver for informed consent. This was a retrospective multicenter cohort study using the Pediatrix CDW. The CDW includes records from approximately 400 community and academic NICUs (levels I–IV) in 35 US states and Puerto Rico, which provided care to approximately 20% of preterm infants born in the United States during the study period. Data was entered by clinicians using admission, progress, and discharge notes, as well as results from laboratory, imaging, and other diagnostic studies. These data were then extracted, consolidated, and processed to populate the CDW.^27^ Data included demographic and maternal characteristics, diagnoses made by clinicians (genetic disorders, morbidities), and hospital disposition (mortality, discharge home, or transfer). The results of negative test results, including for genetic disorders, were not available. Records of 409 399 infants born from 23 to 33 weeks’ gestation, discharged from 2000 to 2020 were extracted. Data from NICU admission to discharge were collected. Birth weight was normalized within the cohort by gestational age and sex. Cases were excluded if these data were missing. Infants were classified as SGA for normalized birth weight below the 10^th^ percentile or as appropriate for gestational age (AGA) at the 10^th^ percentile or above and less than the 90^th^ percentile. Infants with birth weight greater than the 90^th^ percentile were excluded to prevent the confounding impact of poor outcomes in LGA infants. We determined the overlap of this definition with SGA defined by Fenton criteria and by the Eunice Kennedy Shriver National Institute of Child Health and Human Development (NICHD) consensus FGR definition.^10,28^

### Association of Genetic Disorders with SGA versus AGA birth among preterm infants

We performed multiple variable logistic regression of the association of the following maternal and infant characteristics: ethnicity/race (maternal self-declared), maternal age, diagnosis of genetic disorders, the presence of congenital anomalies (defined as clinician diagnosed anomalies of the heart, gastrointestinal tract, brain, lungs, kidneys and urinary tract, or the presence of hydrops), and year of discharge. Congenital anomalies themselves are strongly associated with genetic disorders. Therefore, we tested for an interaction between congenital anomalies and genetic disorders with the outcome of SGA birth.

This analysis was also repeated limiting the cohort to infants born following singleton gestation to test the effect of multiple gestation. Unadjusted odds of SGA versus AGA birth were determined for each genetic disorder in which SGA birth was observed. The prevalence rate of SGA birth across maternal age was also determined.

### Association of genetic disorders with morbidity and mortality within preterm SGA infants

Next, we evaluated factors associated with severe morbidity or mortality within preterm SGA infants. To limit ascertainment bias, infants transferred after birth or prior to discharge were excluded from the analysis of clinical outcomes.^29^ Multivariable logistic regression was performed for severe morbidity or mortality as a combined clinical outcome. Severe morbidity was defined as any of the following: acute kidney injury (AKI) defined by the diagnosis of oliguria or anuria (ICD-9 788.5 or ICD-10: R34); severe intracranial hemorrhage (ICH) defined as grade III or IV intraventricular hemorrhage by Papile criteria or cystic periventricular leukomalacia;^30^ medically or surgically treated necrotizing enterocolitis (NEC); severe bronchopulmonary dysplasia (BPD) defined as invasive mechanical ventilation at 36 weeks’ postmenstrual age; severe retinopathy of prematurity (ROP) defined as need for any medical or surgical intervention; culture-positive sepsis defined by any positive blood or urine culture; or shock defined as administration of any vasopressor or inotrope. The following were covariates: exposure to antenatal betamethasone, mode of delivery (vaginal delivery was treated as reference), phenotypic sex (female was treated as reference given the associated risk of morbidity in male infants),^31,32^ race (White race was treated as reference given thee proportional size), gestational age, birth weight Z-score, discharge year epoch (2000-2004, 2005-2009, 2010-2014, 2015-2020), intubation in the first 72 hours of life, presence of a known genetic disorder, and the presence of congenital anomalies. This analysis was repeated limiting the cohort to infants born following singleton gestation to test the effect of multiple gestations. As before, we also tested for an interaction between genetic disorders and congenital anomalies and the outcome of morbidity or mortality.

Finally, we sought to evaluate how SGA preterm infants with or without congenital anomalies compared to AGA preterm infants with respect to rates of morbidity and mortality. Using AGA preterm infants as reference for both analyses, we determined the crude odds ratios (OR) of individual comorbidities for preterm infants born SGA without congenital anomalies and for preterm infants born SGA with congenital anomalies. Analyses were made using RStudio version 2022.07.0 (R Project for Statistical Computing) (eMethods in the **Supplement**).^33^

## Results

378 887 SGA and AGA infants were identified in this cohort (**Table 1**). Of these, 176 791 (46.7%) were female. The mean (SD) gestational age was 30.1 (2.86) weeks, and the mean (SD) birth weight was 1.44 (0.49) kg. Genetic disorders were diagnosed in 5 386 (1.4%) infants. Congenital anomalies were present in 24 463 (6.4%) of infants. 36 985 (9.8%) infants were defined as SGA. 36 985 infants met this definition, all of whom were below the 10^th^ percentile for NICHD fetal growth (**Supplemental Figure 1**). 33 451 (92.1%) of these infants were also below the Fenton 10^th^ percentile definition of SGA birth.

**Table 1.**
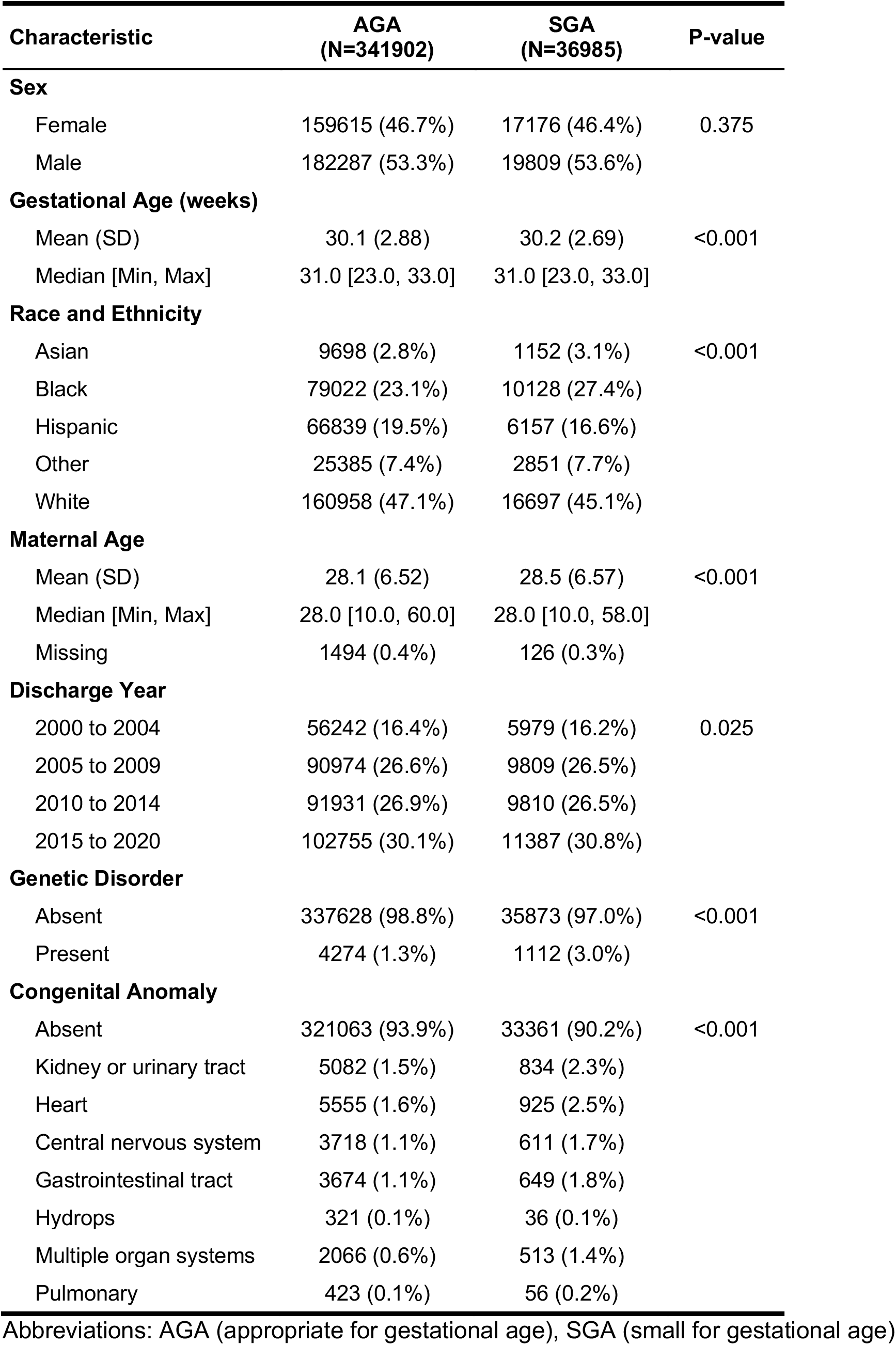
Descriptive characteristics of cohort.

### Association of Genetic Disorders with SGA versus AGA birth among preterm infants

Genetic disorders were present in 1112 (3.0%) of the 36 985 preterm SGA infants and in 4274 (1.3%) of the 341 902 preterm AGA infants in this cohort. Multivariable logistic regression demonstrated higher adjusted OR of SGA versus AGA birth for infants born to mothers identifying as Asian or Black (versus White) and lower for those born to mothers identifying as Hispanic (versus White) (**Figure 1**). Known genetic disorders and congenital anomalies (except for fetal hydrops and pulmonary anomalies) were also found to be associated with SGA birth in preterm infants. SGA birth was not found to vary across the period of this study (2000–2020). Nearly identical results were found for all covariates when this regression was limited to infants born following singleton gestation (**Supplemental Figure 2**). Greater maternal age was also associated with SGA birth (**Figure 3**). The association between genetic disorders and SGA birth was not driven by a single genetic disorder. Rather, SGA birth was associated with multiple known diagnoses including trisomy 13, trisomy 18, trisomy 21, Turner syndrome, unspecified aneuploidy, unspecified copy number variant, sickle cell anemia, cystic fibrosis, hypophosphatasia, and the presence of multiple genetic disorders (**Supplemental Table 1**). As a secondary analysis, we repeated this regression testing for an interaction between the presence of genetic disorders and the presence of any congenital anomaly. We found that these factors each independently associated with SGA versus AGA birth in preterm infants, but we did not find a significant interaction between these factors and SGA birth.

**Figure 1:**
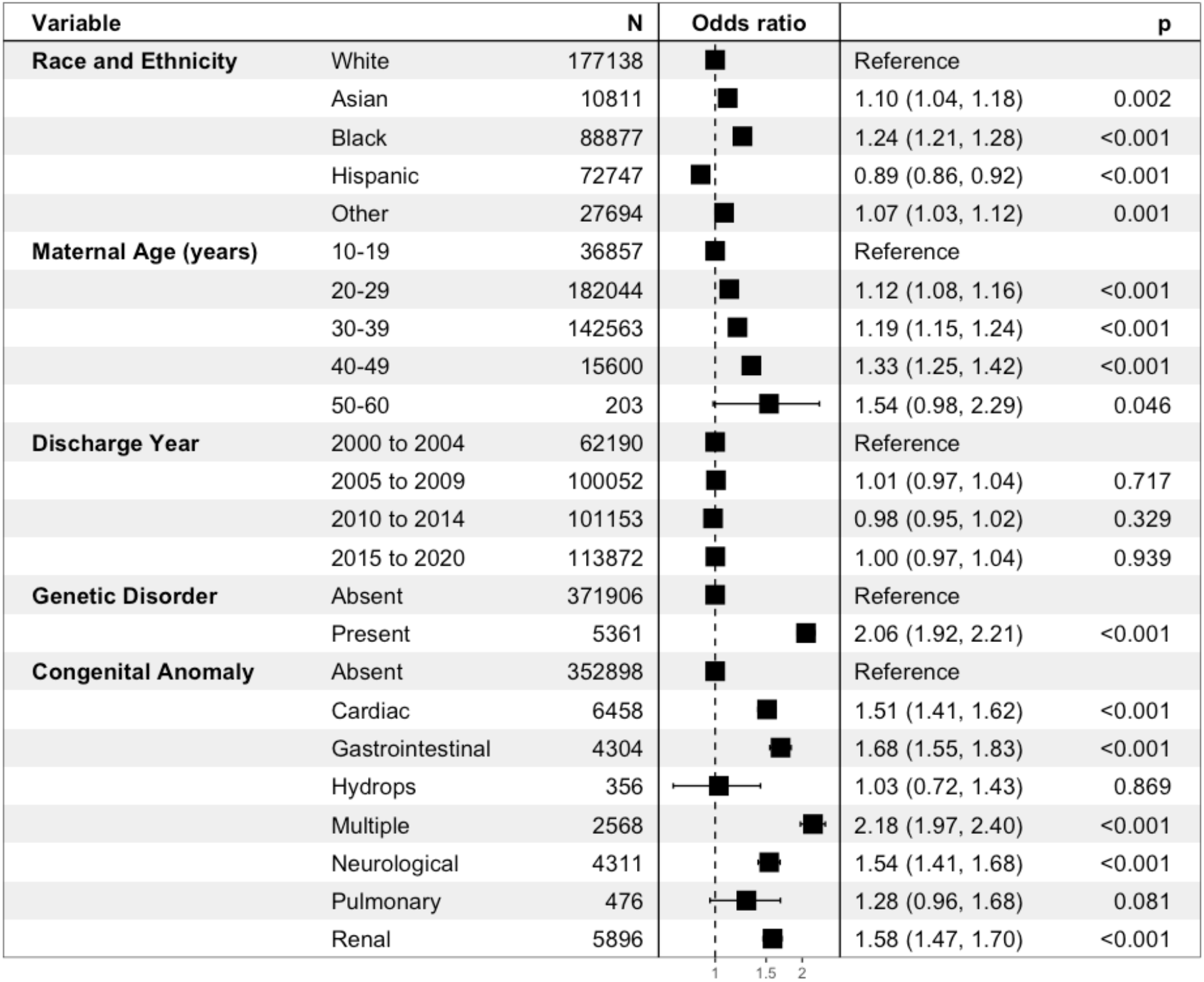
Multivariable regression of clinical characteristics of preterm infants born SGA versus AGA. Regression was performed for the outcome of SGA birth within infants born before 34 weeks’ gestation. Maternal identification as Hispanic was associated with lower adjusted odds of SGA status. Maternal identification as Asian or Black, advancing maternal age, the presence of a known genetic disorder, and the presence of congenital anomalies (excluding hydrops and pulmonary anomalies) were associated with greater adjusted odds of SGA status.

**Figure 2.**
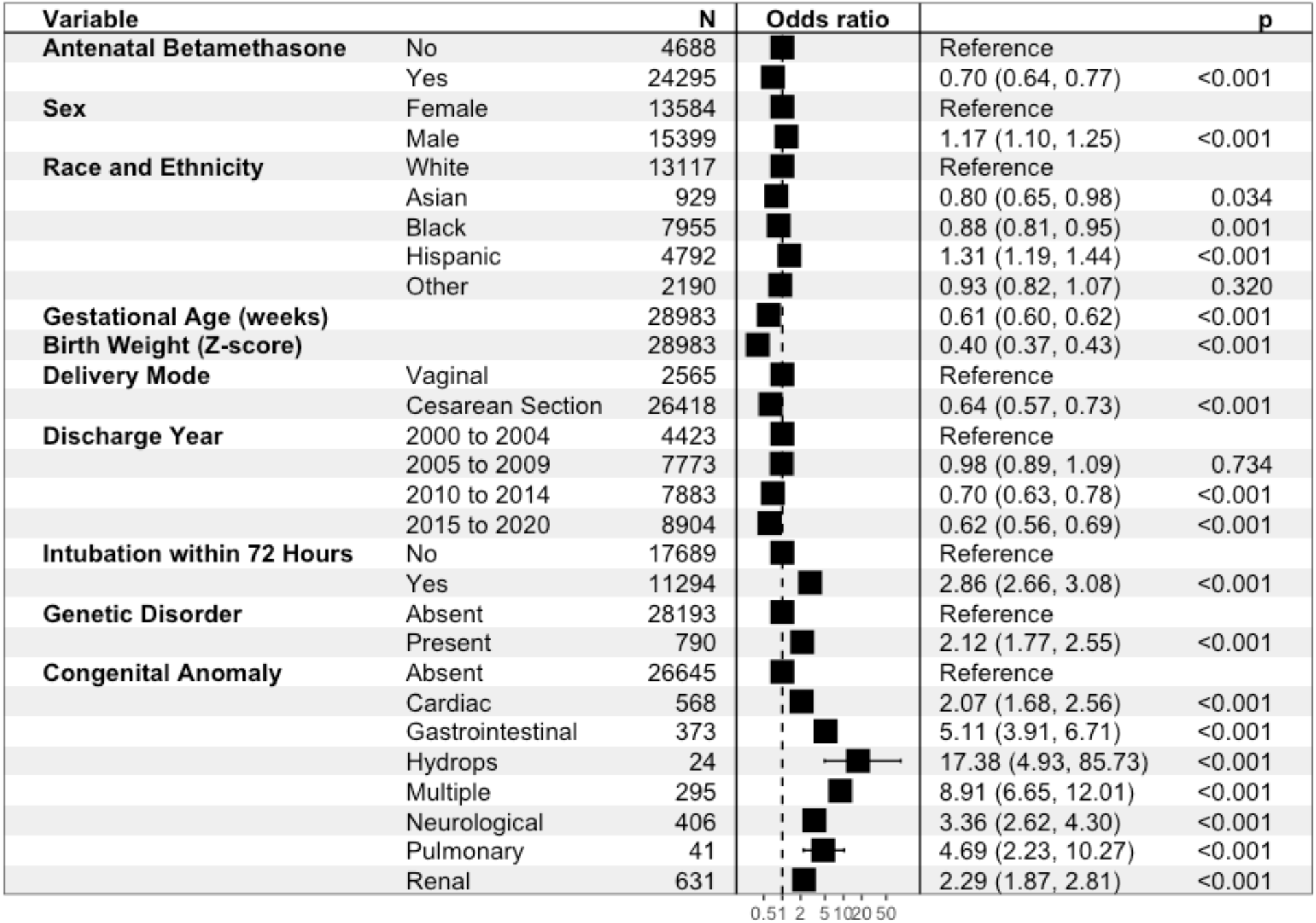
Multivariable logistic regression of clinical characteristics of preterm SGA infants with severe morbidity or mortality (versus preterm SGA infants without severe morbidity or mortality). Within the subset of preterm infants born SGA, regression was performed against the combined outcome of mortality or severe morbidity. Antenatal betamethasone, maternal identification as Asian or Black, increasing gestational age, increasing birth weight, cesarean delivery, and later epochs were found to be associated with lower adjusted odds of the composite outcome. Male sex, maternal identification as Hispanic, intubation after birth, and the presence of known genetic disorders or congenital anomalies were associated with greater adjusted odds of the composite outcome.

**Figure 3.**
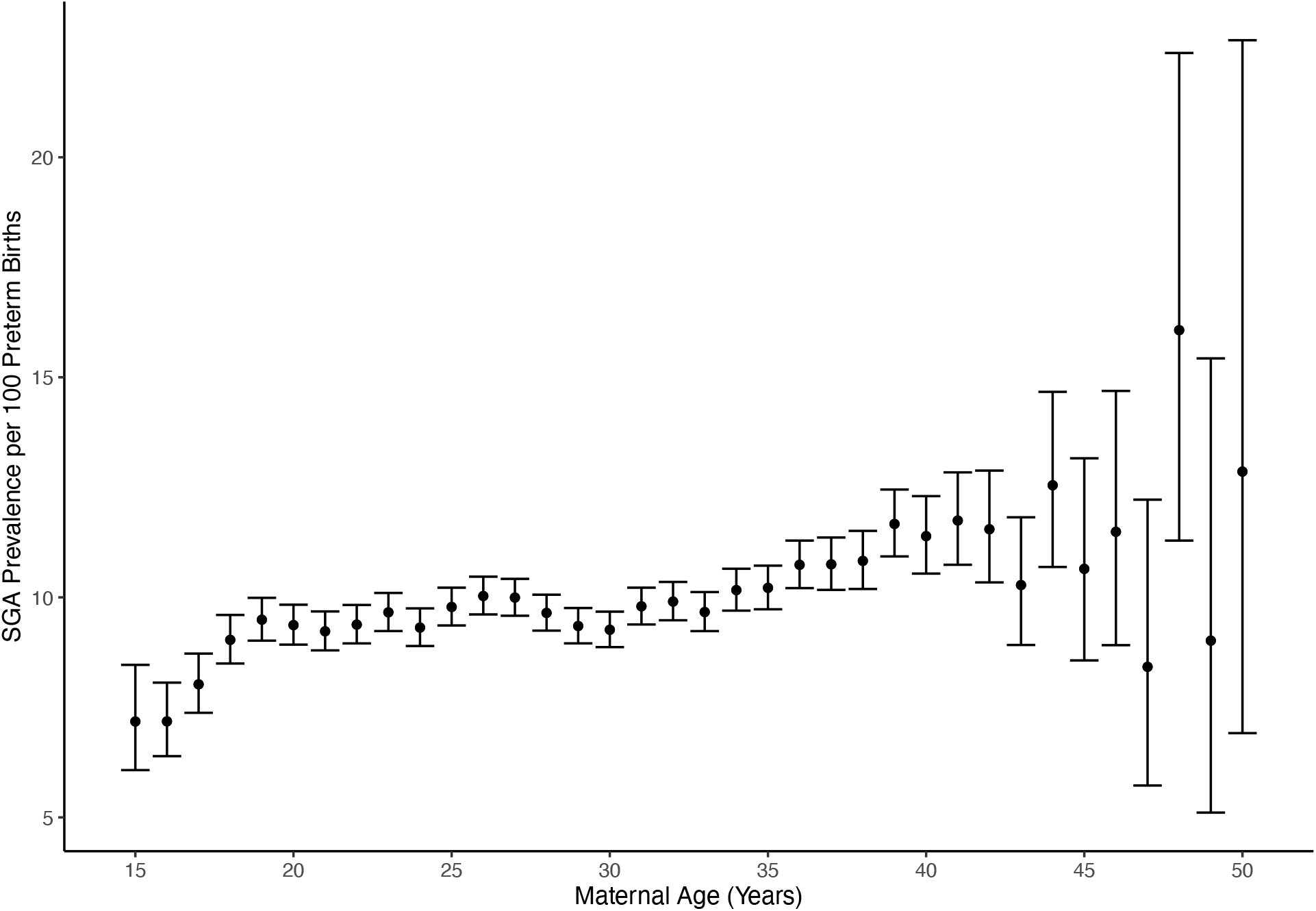
Prevalence of SGA birth by maternal age. The prevalence of SGA birth was determined (within preterm births) for each year of maternal age from 15 to 50. Error bars depict the 95% CI. Extreme maternal ages, less than 15 and greater than 50, were excluded from the analysis due to their small sample size. For each one year increase in maternal age, on average, there was a 0.107 increase in SGA prevalence per 100 preterm births (adjusted r^2^ = 0.46, p = <0.001).

### Association of genetic disorders with morbidity and mortality within preterm SGA infants

We next focused on morbidity and mortality within the subset of 36 985 SGA preterm infants. After excluding infants transferred in or to other facilities, and those with missing data, 28 983 SGA preterm infants remained (**Figure 2**). Genetic disorders were present in 375 (4.3%) of the 8810 preterm SGA infants that experienced severe morbidity or mortality as compared to 415 (2.1%) of the 20 173 preterm AGA infants that did not experience these outcomes. Antenatal betamethasone, birth in later years, maternal identification as Asian or Black (versus White), increased gestational age, and increased birth weight all were associated with lower adjusted odds of morbidity or mortality. Male sex, maternal identification as Hispanic (versus White), intubation in the first 72 hours of life, the presence of known genetic disorders, or congenital anomalies were associated with greater adjusted odds of morbidity or mortality. Nearly identical results were found for all covariates when this regression was limited to infants born following singleton gestation pregnancy (**Supplemental Figure 3**). Compared to AGA preterm infants, SGA preterm infants with or without other congenital anomalies experienced higher rates of death and every comorbidity, with the exception of severe ICH, which was observed to occur less frequently in infants with isolated SGA status compared to all AGA infants. (**Table 2**). As a secondary analysis, we repeated this regression with a test for an interaction between the presence of genetic disorders and the presence of any congenital anomaly. We found that SGA preterm infants with genetic disorders and congenital anomalies had an adjusted odds ratio (95% confidence interval) of 1.52 (1.03, 2.27) for severe morbidity or mortality.

**Table 2.** Unadjusted odds of mortality or severe morbidities in preterm SGA infants.

| Outcome | OR (99.44% CI) |  |
| --- | --- | --- |
|  | Preterm SGA Infants without Congenital Anomalies (versus AGA Preterm Infants) | Preterm SGA Infants with Additional Congenital Anomalies (versus AGA Preterm Infants) |
| Mortality or any morbidity | 1.45 (1.39, 1.50) | 4.86 (4.33, 5.46) |
| Mortality | 1.99 (1.87, 2.12) | 5.96 (5.20, 6.83) |
| Acute kidney injury | 1.60 (1.46, 1.75) | 5.03 (4.22, 6.00) |
| Severe intracranial hemorrhage | 0.82 (0.75, 0.91) | 1.89 (1.52, 2.36) |
| Severe necrotizing enterocolitis | 1.28 (1.18, 1.40) | 3.50 (2.91, 4.21) |
| Severe bronchopulmonary dysplasia | 2.40 (2.13, 2.70) | 9.50 (7.78, 11.59) |
| Severe retinopathy of prematurity | 1.52 (1.33, 1.74) | 3.14 (2.31, 4.27) |
| Sepsis | 1.37 (1.29, 1.45) | 3.22 (2.81, 3.68) |
| Shock | 1.35 (1.28, 1.43) | 3.18 (2.78, 3.64) |

321 Abbreviations: CI (confidence interval), OR (odds ratio), SGA (small for gestational age)

## Discussion

The primary finding of our study was that genetic disorders were strongly and nonspecifically associated with preterm SGA birth, morbidity, and mortality. There are important strengths, limitations, and clinical implications of this finding. Our analysis revealed several secondary findings as well: lower birth weight and higher risk of illness in preterm infants with cystic fibrosis and hypophosphatasia, a synergistic risk of illness in infants with congenital anomalies and genetic disorders, and a lower rate of severe ICH in preterm SGA infants without congenital anomalies. In the following we discuss the primary and secondary findings of our research. Using a highly generalizable cohort, we found that commonly diagnosed genetic disorders were twice as prevalent in SGA infants compared to AGA infants, and that genetic disorders were strongly associated with SGA birth weight after adjusting for known risk factors. Genetic disorders were also strongly associated with severe morbidity and mortality in this population. We confirmed previously described associations between common aneuploidies and SGA birth, morbidity, and mortality. Interestingly, we found a broad association between genetic disorders and preterm SGA birth. Of the 20 genetic disorders analyzed (including “unspecified” cases of aneuploidy or copy number variants), significant associations with SGA birth were found in 10, and nonsignificant trends towards this association were found in 8. Cystic fibrosis has recently been implicated as a cause of lower birth weight and earlier gestation of birth,^34^ which our data support. We also found evidence of a novel association between hypophosphatasia and preterm SGA birth, as well as significantly greater odds of morbidity or mortality. If confirmed, these findings might illuminate pathogenic mechanisms of growth restriction.

Multiple gestations have typically been excluded in large cohort studies limiting the generalizability of findings. We found similar results with or without this exclusion. However, we did not evaluate pairwise outcomes within twin or greater multiple gestations. Clinicians and families, particularly in the setting of growth discordant twins, often must consider risks and benefits in timing of delivery. Targeted focus on SGA multiple gestations could help better inform these decisions, as well as validate twin-specific growth charts.^35^ Unique factors in twin pregnancies including unequal placentation, twin-twin transfusion, and birth weight discordance all may influence clinical outcomes, and should be explored further.^36,37^

In addition to genetic disorders, we found that congenital anomalies were strongly associated with preterm SGA birth, morbidity, and mortality. Congenital anomalies of the kidney, heart, brain, and other organ systems are themselves strongly linked to genetic disorders. It is unclear if anomalies themselves caused lower birth weight and illness, or if anomalies were the sentinel features of underlying genetic disorders. A parsimonious explanation is that underlying genetic disorders contribute to congenital anomalies, poor fetal growth, and severe morbidity. This is further supported by the finding that genetic disorders and congenital anomalies interacted to confer synergistically greater risk of morbidity and mortality in SGA preterm infants.

Intriguingly, preterm SGA infants without congenital anomalies were found to have lower odds of severe ICH compared to AGA infants. Fetal growth restriction and SGA birth have been described as protective against severe intraventricular hemorrhage in extremely preterm (born < 28 weeks’ gestation) infants.^6,8,38–40^ The implication of a lower risk of severe ICH in SGA birth without other anomalies warrants attention given reports of higher risk of neurodevelopmental impairment including poorer cognitive outcomes in this population.^41^ Administration of antenatal corticosteroids reduces the risk of severe IVH and mortality, but studies have suggested that steroids in SGA preterm infants may not reduce serious morbidities.^42,43^ Although a previous landmark trial reported a potential reduction in severe IVH but no effect on neurosensory impairment with the use of prophylactic indomethacin in extremely low birth weight infants, multiple studies have refuted this finding.^44–46^ Replication, functional validation, and basic investigation into protection of IVH in growth restricted extremely preterm infants may inform novel therapies.

Our study had several compelling strengths. This represents the largest and most comprehensive examination of factors associated with preterm SGA status and comorbidities to our knowledge. This was a multicenter investigation of infants from diverse backgrounds, with generalizable results. There are important limitations to this study. Infants born SGA may be more likely to undergo pre- or postnatal genetic testing, especially in the presence of other congenital anomalies or comorbidities. Details regarding testing, including time (pre- or postnatal) and indication are unknown in this cohort. Genetic testing has also undergone dramatic changes from 2000 to 2020. There is likely substantial heterogeneity in the genetic workups performed across this period. Unfortunately, data are not available regarding the spectrum of testing performed in the cohort, including negative results. There may also be heterogeneity in the assignment of gestational age. The method used to determine this for each individual in the cohort is unknown. Additionally, SGA and FGR are defined differently in the literature of this field. We defined SGA birth here using the distribution of birth weight found within this cohort. This definition increases the internal validity or our analysis but may limit generalization. Given that our definition of SGA shared 92.1% with Fenton-defined SGA infants, this is unlikely. The optimal criteria for defining a newborn as SGA or growth restricted likely include individual environmental, genetic, and biological factors, many of which are yet unknown. These analyses also did not consider when growth restriction occurred during pregnancy, patterns of growth restriction (such as head-sparing or symmetric), fetal exposures, postnatal growth patterns, or social determinants of health. Our dataset lacks maternal records, including the presence of comorbidities that may contribute to placental insufficiency. Our finding that extremes of maternal age, as well as increasing maternal age from 18 to 34 years, were associated with SGA status likely reflects the contribution of these maternal factors.

Genetic counseling and consideration of testing for FGR is recommended by the American College of Obstetricians and Gynecologists as well as by the Society for Maternal and Fetal Medicine.^47,48^ There is not yet consensus regarding the postnatal evaluation of SGA infants.^49,50^ Scarce data exist regarding how often genetic workups or testing are actually performed pre- or postnatally for these indications. Furthermore, perinatal genetic disorders often present nonspecifically and are often diagnosed belatedly.^51,52^ The indications and potential benefit for genetic testing in this population is unknown without prospective, genome-wide testing.

Our findings have implications for clinical practice and further research. Clinicians should strongly consider genetic testing of preterm SGA infants, particularly in the setting of other comorbidities or anomalies. Genetic disorders were found in 3.0% of preterm SGA infants overall and 4.3% of preterm SGA infants with severe morbidity and mortality. These represent underestimates as prospective testing was not preformed. Without prospective genetic testing the baseline rate genetic disorders in this population is unknown. Preterm SGA infants with genetic disorders or congenital anomalies may be at higher risk for morbidity and mortality. Clinicians should consider this risk when counseling families, evaluating the risks and benefits of management decisions, or estimating the pre-test probability of diagnostic tests. Prospective genomic research is urgently needed to clarify the contribution of genetic disorders to disease in this population.

## Data Availability

All data produced in the present study are available upon reasonable request to the authors

## Acknowledgements

The authors wish to gratefully acknowledge Dr. Ali G. Gharavi (Columbia University Irving Medical Center) and his research team, as well as Dr. Joseph M. Feinglass (Feinberg School of Medicine, Northwestern University) for their mentorship and thoughtful review of this project.

Authors TH and JEM are supported by *Thrasher Research Fund Early Career Awards.* TH is also supported by the *NIH National Center for Advancing Translational Sciences* (KL2TR001874).

## Supplemental Materials

**Supplemental Table 1.** Unadjusted odds of SGA birth within diagnosed genetic disorders.

**Supplemental Figure 1.** Classification of SGA or FGR birth in this cohort by related criteria.

**Supplemental Figure 2.** Multivariable logistic regression for SGA versus AGA status in preterm infants with singleton gestation.

**Supplemental Figure 3.** Multivariable logistic regression for severe morbidity or mortality in SGA preterm infants following singleton gestation.

**eMethods.** R studio script used for this project.

---

**Supplemental Table 1.** Unadjusted odds of SGA birth associated for specific genetic disorders.

| Genetic Disorder | SGA (n) | AGA (n) | OR (99.75% CI) |
| --- | --- | --- | --- |
| **Aneuploidy** |  |  |  |
| Trisomy 13 | 32 | 109 | 2.76 (1.50–5.08) |
| Trisomy 18 | 145 | 80 | 17.06 (11.19-26.00) |
| Trisomy 21 | 286 | 987 | 2.73 (2.22-3.34) |
| Trisomy 22 | 1 | 2 | 4.71 (0.12-190.89) |
| Turner syndrome | 17 | 34 | 4.71 (1.92-11.55) |
| Klinefelter syndrome | 9 | 35 | 2.42 (0.78-7.49) |
| Unspecified aneuploidy | 204 | 554 | 3.47 (2.70-4.44) |
| **Copy Number Variant** |  |  |  |
| 13q deletion | 3 | 8 | 3.53 (0.46-27.33) |
| Cri-du-chat | 4 | 17 | 2.21 (0.41-11.88) |
| DiGeorge syndrome | 9 | 54 | 1.57 (0.53-4.66) |
| Unspecified copy number variant | 15 | 31 | 4.55 (1.76-11.79) |
| **Hematologic Single Gene Disorder** |  |  |  |
| Hemophilia A | 1 | 12 | 0.78 (0.03-18.25) |
| Thalassemia | 169 | 1256 | 1.27 (0.99-1.62) |
| Hemoglobin Bart's | 39 | 391 | 0.94 (0.56-1.56) |
| Sickle cell anemia | 48 | 196 | 2.30 (1.42-3.75) |
| G6PD deficiency | 32 | 200 | 1.51 (0.85-2.68) |
| Fanconi anemia | 2 | 1 | 18.82 (0.46-763.54) |
| **Miscellaneous** |  |  |  |
| Cystic fibrosis | 48 | 206 | 2.19 (1.35-3.56) |
| Hypophosphatasia | 41 | 80 | 4.82 (2.70-8.62) |
| Multiple diagnoses | 7 | 9 | 7.32 (1.60-33.59) |

Abbreviations: AGA (appropriate for gestational age), CI (confidence interval), OR (odds ratio), SGA (small for gestational age)

**Supplemental Figure 1.**
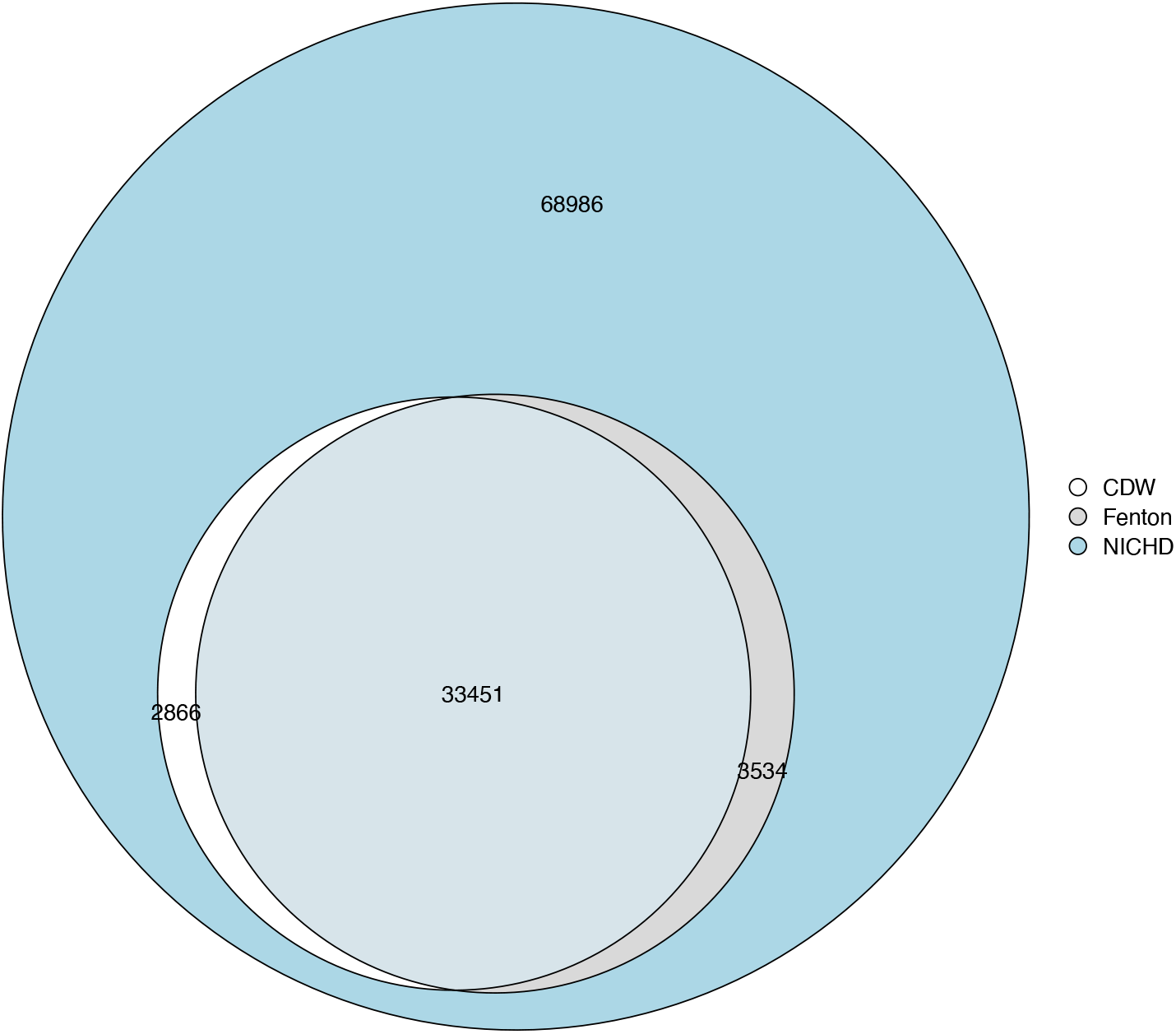
Classification of SGA or FGR birth in this cohort by related criteria. This cohort included 409,399 preterm infants from the Pediatrix CDW. SGA was defined for this study as below the 10^th^ percentile for gestational age and sex within the CDW cohort. 36,985 infants met this definition, all of whom were below the 10^th^ percentile for NICHD fetal growth. 33,451 (92.1%) of these infants also fell below the Fenton 10^th^ percentile definition of SGA birth. The numbers of infants meeting these criteria are depicted in the Euler diagram above.

**Supplemental Figure 2.**
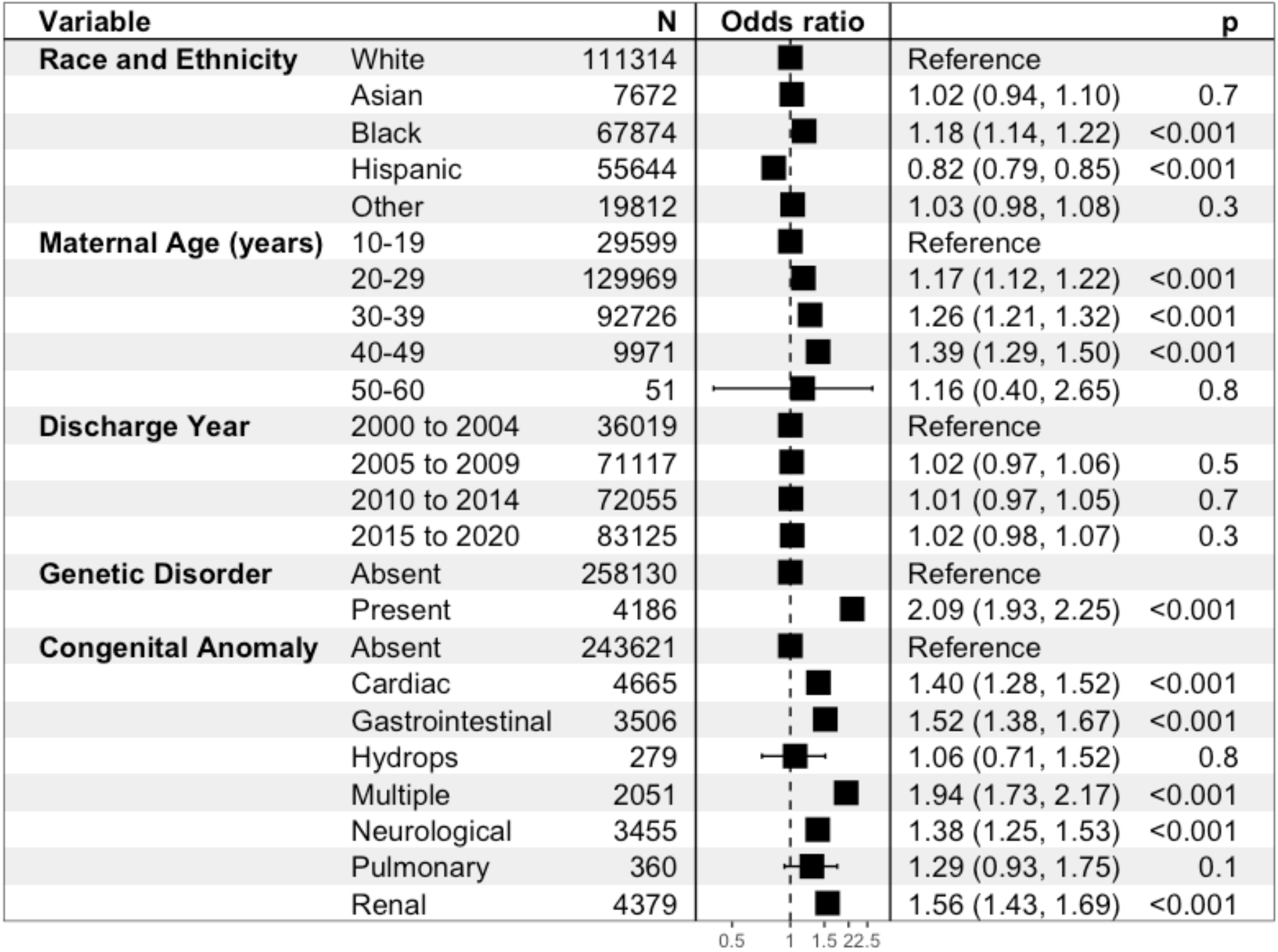
Multivariable logistic regression for SGA versus AGA status in preterm infants with singleton gestation. The outcome of preterm SGA status was modeled as before (Figure 1) in only infants born following singleton gestation. Nearly identical adjusted odds were found for all covariates.

**Supplemental Figure 3.**
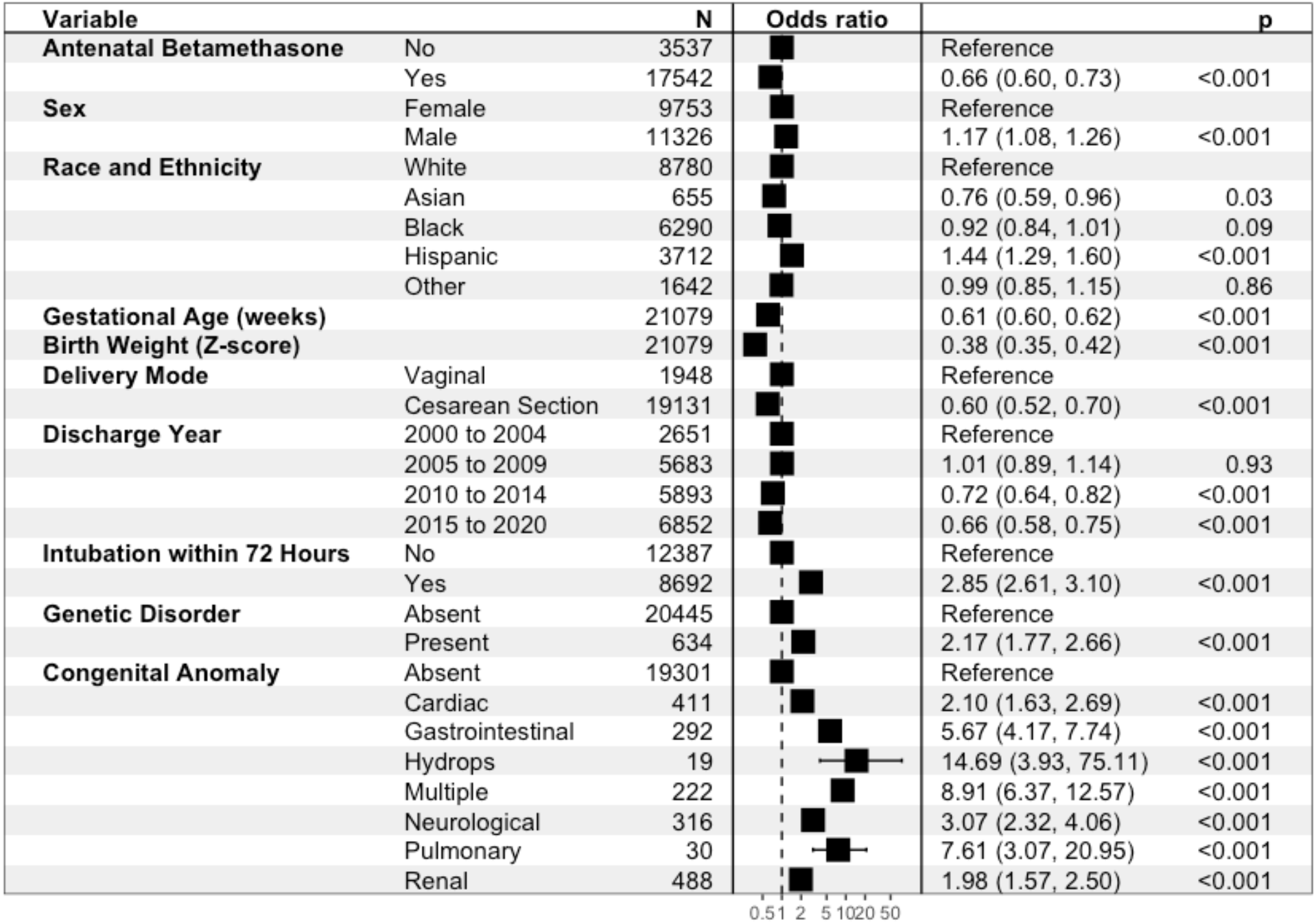
Multivariable logistic regression for severe morbidity or mortality in SGA preterm infants following singleton gestation. The combined outcome of mortality or severe morbidity status for preterm SGA infants was modeled as before (Figure 2) in only infants born following singleton gestation. Nearly identical adjusted odds were found for all covariates.

## eMethods

The following script was used to generate tables and figures. The script was created using RStudio (version 2022.07.0).

**#### Table 1**

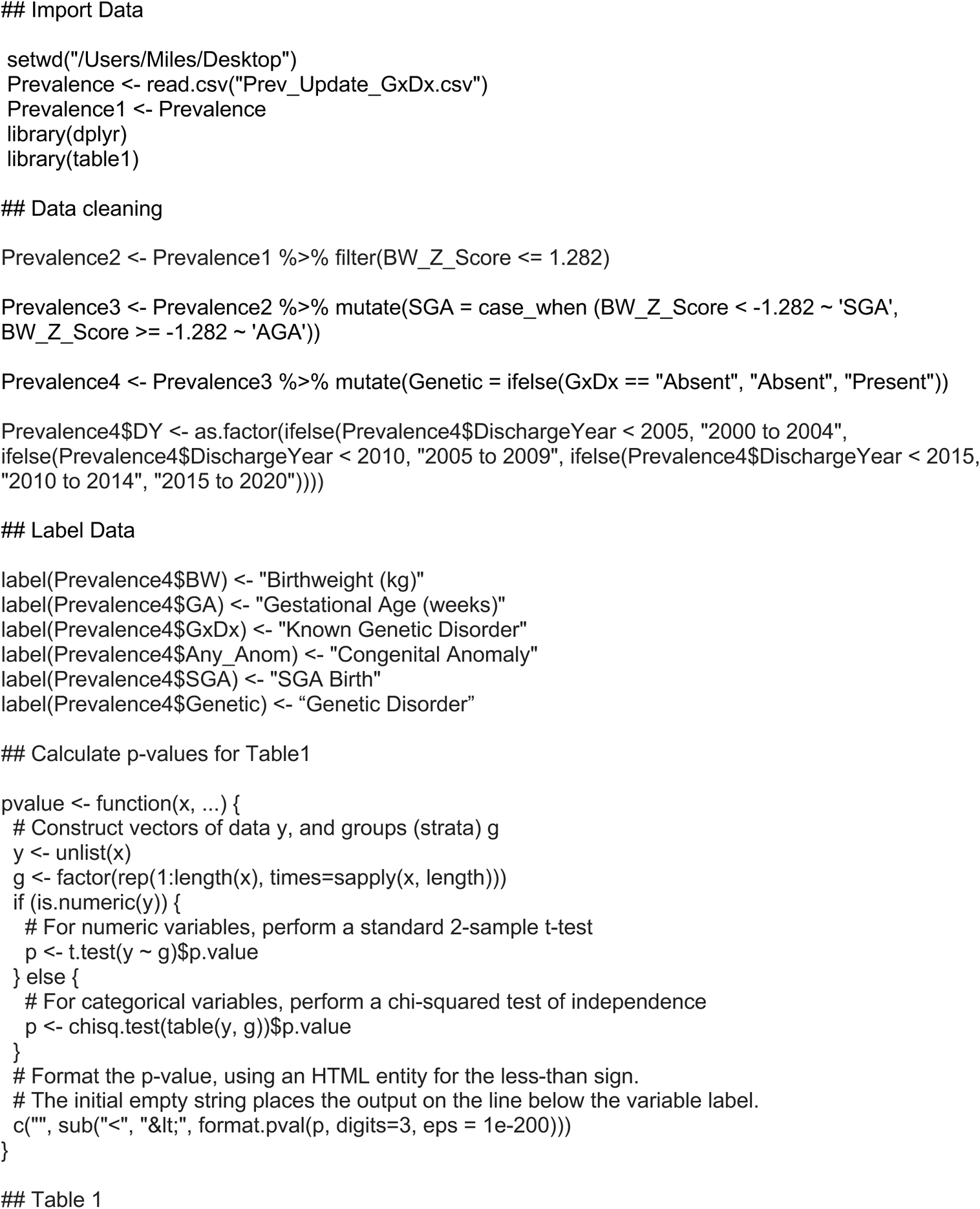
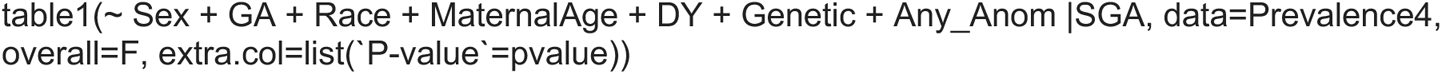

**####Table 2:**

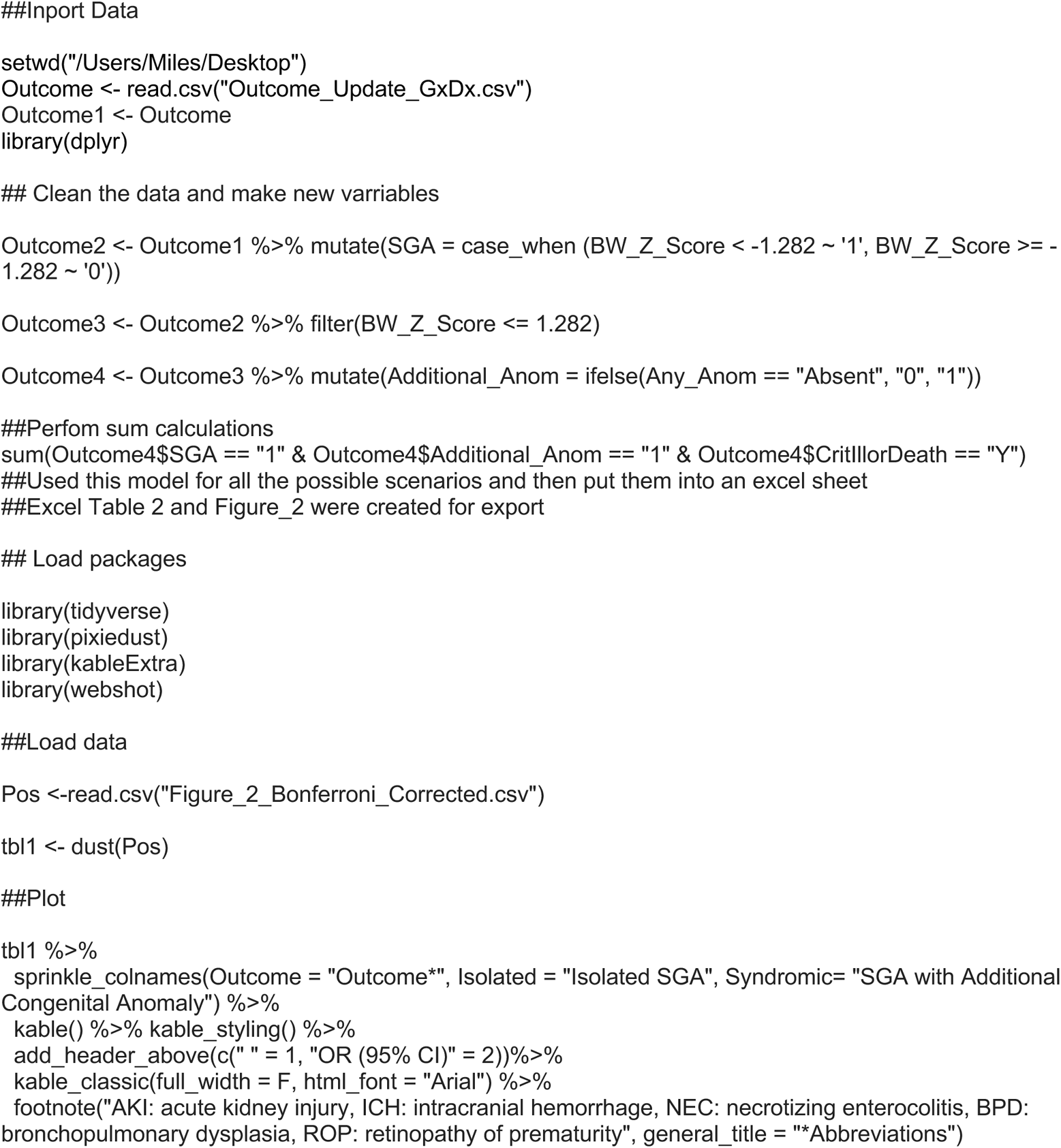

**####Figure 1**

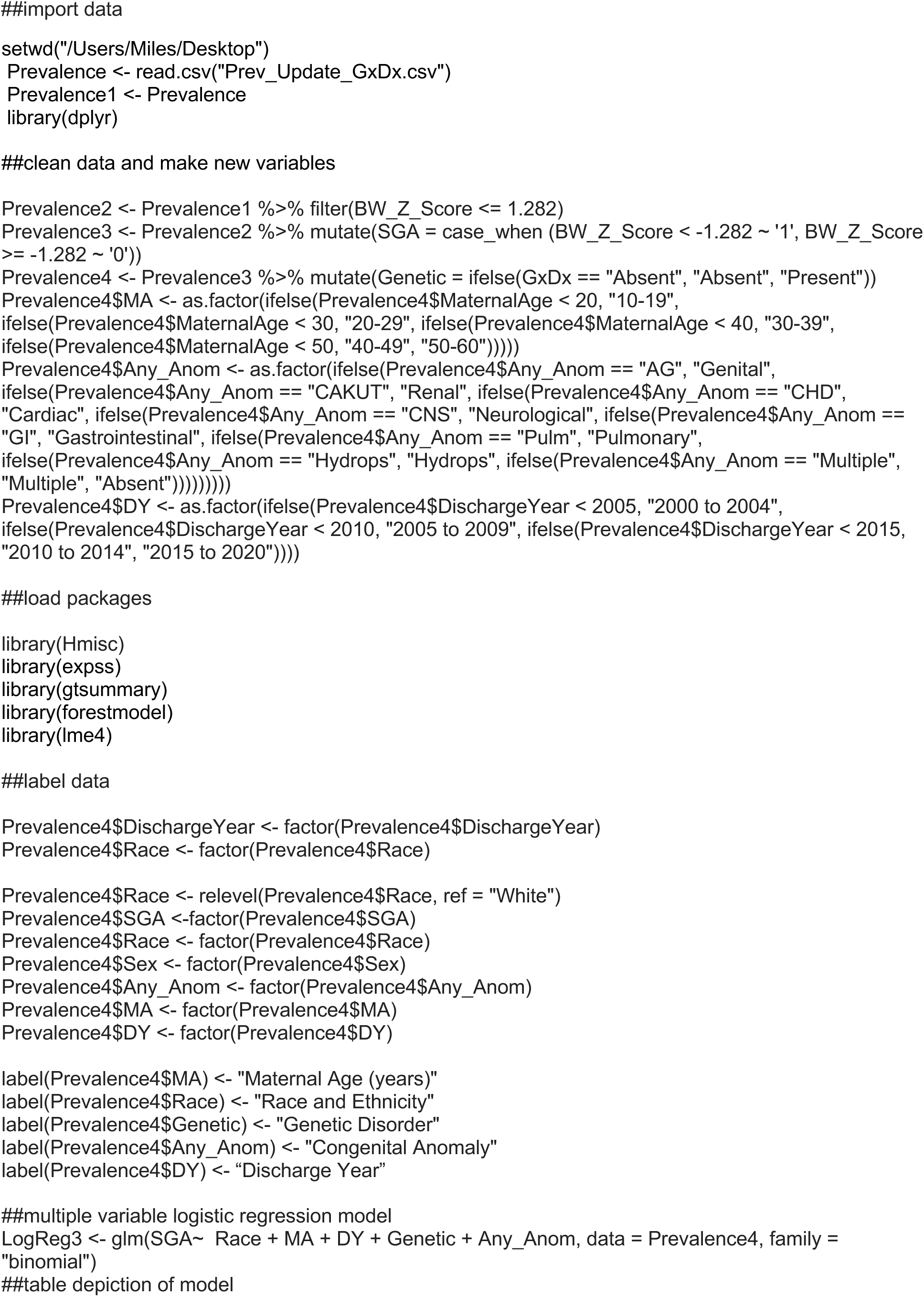
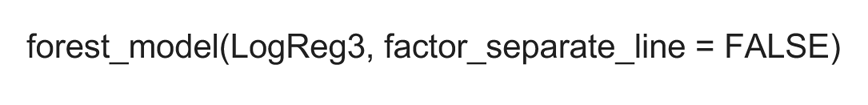

**####Figure 2:**

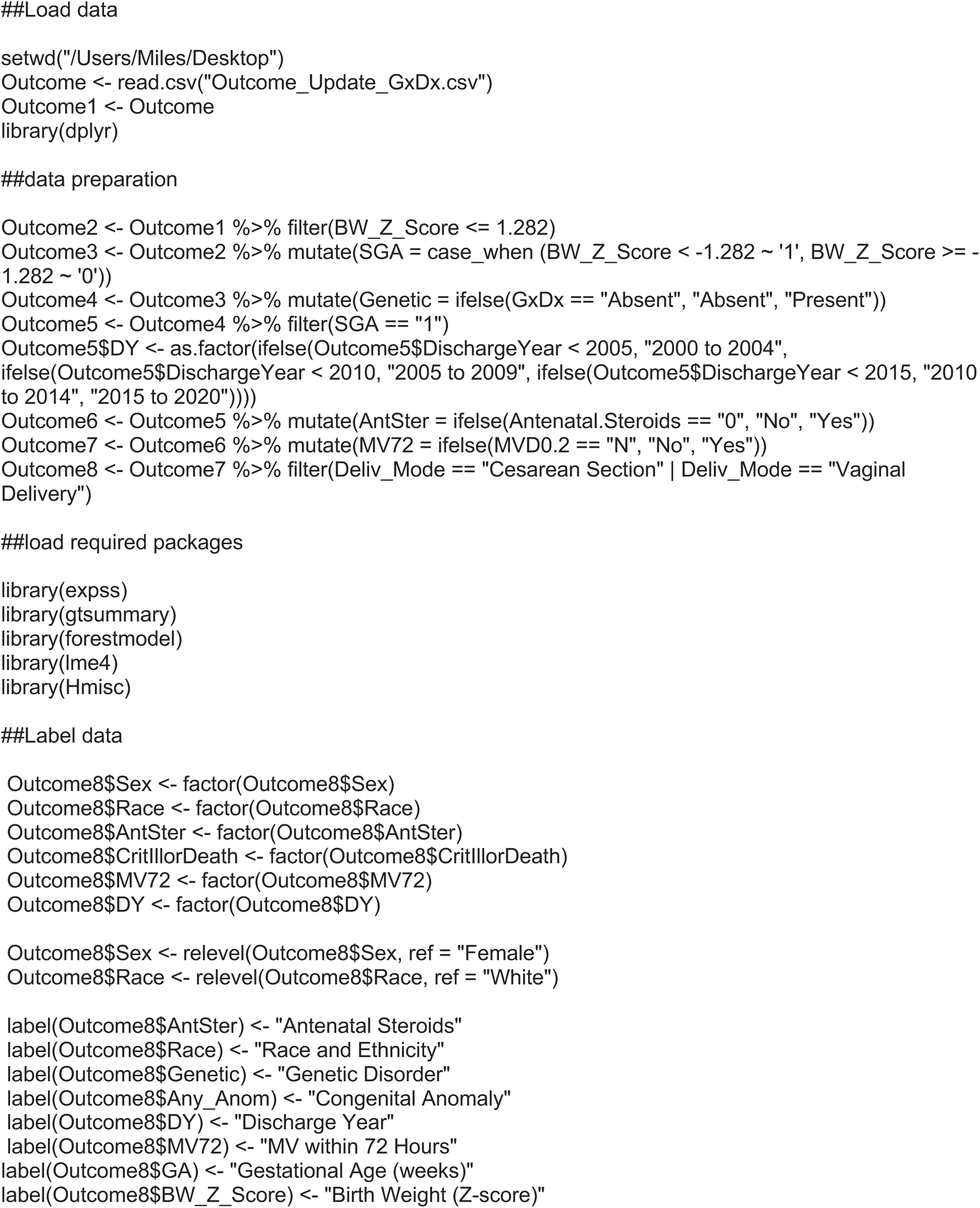
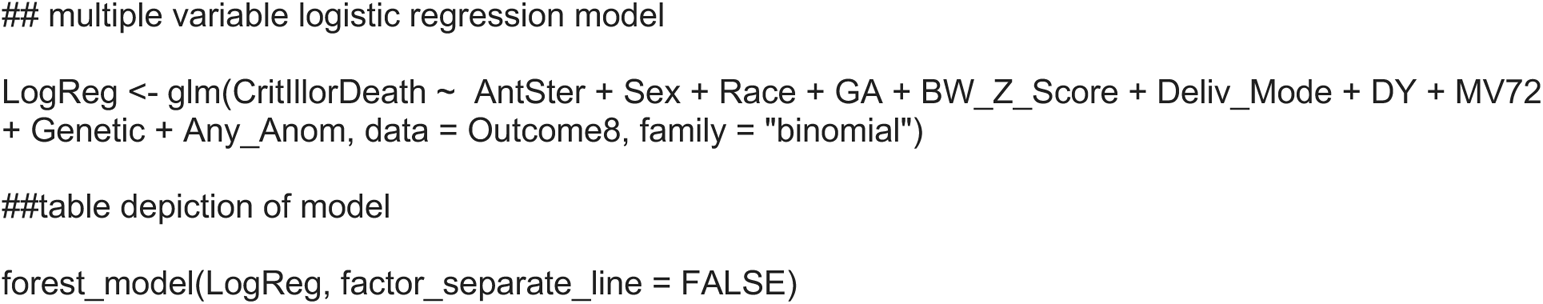

**####Figure 3:**

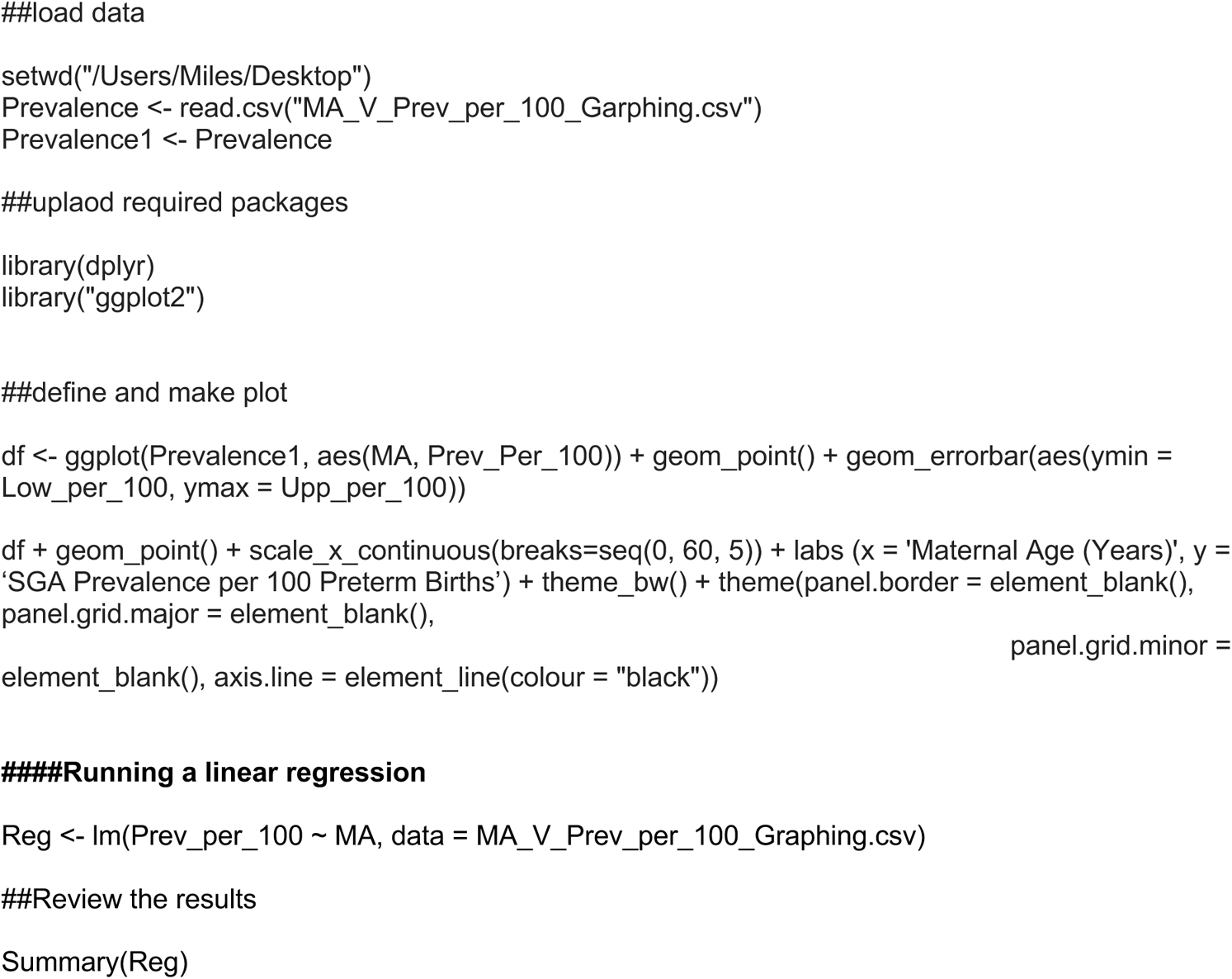

**####Supplemental table 1**

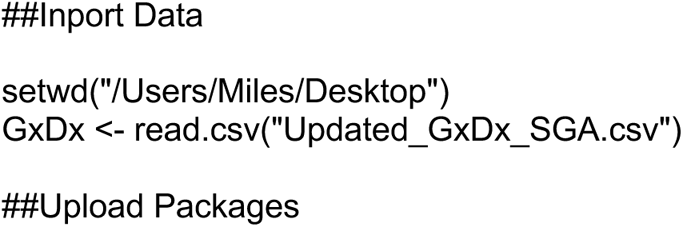
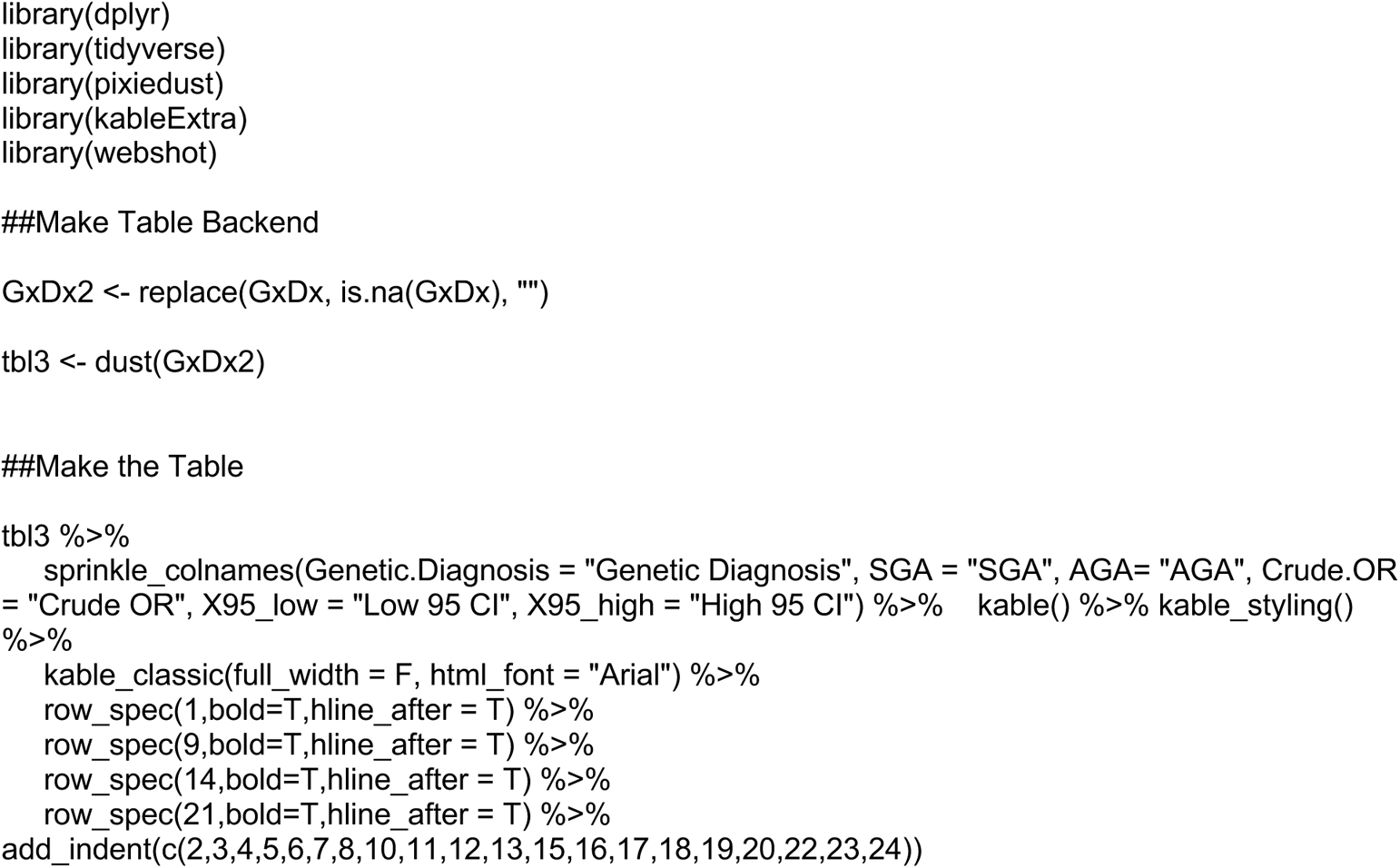

**####Supplemental Figure 1**

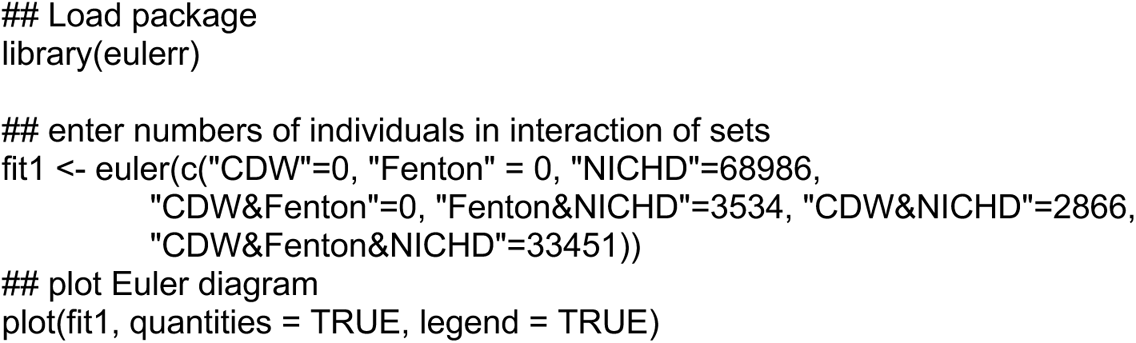

**####Supplemental Figure 2**

##Same code as Figure 1 but added one line of code in data preparation

Prevalence4 <- Prevalence4 %>% filter(BirthNumber == “1”)

**####Supplemental Figure 3**

##Same code as Figure 2 but added one line of code in data preparation

Outcome8 <- Outcome8 %>% filter(BirthNumber == “1”)

**####Figure 1 Interaction Test**

LogReg3 <- glm(SGA~ Race + MA + DY + Genetic + Any_Anom + Genetic*Any_Anom, data =

Prevalence4, family = “binomial”)

anova(LogReg3, test=’Chisq’)

summary(LogReg3)

**#### Interaction Test**

LogReg5 <- glm(CritIllorDeath ~ Antenatal.Steroids + Sex + Race + GA + MVD0.2 + Any_Anom + DY + BW_Z_Score + Genetic + Deliv_Mode + Any_Anom + Genetic*Any_Anom, data = Outcome8,

family = “binomial”)

anova(LogReg5, test=’Chisq’)

summary(LogReg5)

## References

1. Bernstein IM, Horbar JD, Badger GJ, Ohlsson A, Golan A. Morbidity and mortality among very-low-birth-weight neonates with intrauterine growth restriction. The Vermont Oxford Network. Am J Obstet Gynecol. Jan 2000;182(1 Pt 1):198–206. doi:10.1016/s0002-9378(00)70513-8

2. Zeitlin J, El Ayoubi M, Jarreau PH, et al. Impact of fetal growth restriction on mortality and morbidity in a very preterm birth cohort. J Pediatr. Nov 2010;157(5):733–9.e1. doi:10.1016/j.jpeds.2010.05.002

3. Peacock JL, Lo JW, D’Costa W, Calvert S, Marlow N, Greenough A. Respiratory morbidity at follow-up of small-for-gestational-age infants born very prematurely. Pediatr Res. Apr 2013;73(4 Pt 1):457–63. doi:10.1038/pr.2012.201

4. De Jesus LC, Pappas A, Shankaran S, et al. Outcomes of small for gestational age infants born at <27 weeks’ gestation. J Pediatr. Jul 2013;163(1):55–60.e1-3. doi:10.1016/j.jpeds.2012.12.097

5. Garite TJ, Clark R, Thorp JA. Intrauterine growth restriction increases morbidity and mortality among premature neonates. Am J Obstet Gynecol. Aug 2004;191(2):481–7. doi:10.1016/j.ajog.2004.01.036

6. Mendez-Figueroa H, Truong VT, Pedroza C, Chauhan SP. Morbidity and Mortality in Small-for-Gestational-Age Infants: A Secondary Analysis of Nine MFMU Network Studies. Am J Perinatol. Mar 2017;34(4):323–332. doi:10.1055/s-0036-1586502

7. Horbar JD, Carpenter JH, Badger GJ, et al. Mortality and neonatal morbidity among infants 501 to 1500 grams from 2000 to 2009. Pediatrics. Jun 2012;129(6):1019–26. doi:10.1542/peds.2011-3028

8. Baer RJ, Rogers EE, Partridge JC, et al. Population-based risks of mortality and preterm morbidity by gestational age and birth weight. J Perinatol. Nov 2016;36(11):1008–1013. doi:10.1038/jp.2016.118

9. Minor KC, Bianco K, Sie L, Druzin ML, Lee HC, Leonard SA. Severity of small-for-gestational-age and morbidity and mortality among very preterm neonates. J Perinatol. Apr 2023;43(4):437–444. doi:10.1038/s41372-022-01544-w

10. Fenton TR, Kim JH. A systematic review and meta-analysis to revise the Fenton growth chart for preterm infants. BMC Pediatr. Apr 20 2013;13:59. doi:10.1186/1471-2431-13-59

11. Clayton PE, Cianfarani S, Czernichow P, Johannsson G, Rapaport R, Rogol A. Management of the child born small for gestational age through to adulthood: a consensus statement of the International Societies of Pediatric Endocrinology and the Growth Hormone Research Society. J Clin Endocrinol Metab. Mar 2007;92(3):804–10. doi:10.1210/jc.2006-2017

12. Lee PA, Chernausek SD, Hokken-Koelega AC, Czernichow P. International Small for Gestational Age Advisory Board consensus development conference statement: management of short children born small for gestational age, April 24-October 1, 2001. Pediatrics. Jun 2003;111(6 Pt 1):1253–61. doi:10.1542/peds.111.6.1253

13. Katz J, Lee AC, Kozuki N, et al. Mortality risk in preterm and small-for-gestational-age infants in low-income and middle-income countries: a pooled country analysis. Lancet. Aug 3 2013;382(9890):417–425. doi:10.1016/s0140-6736(13)60993-9

14. Finken MJJ, van der Steen M, Smeets CCJ, et al. Children Born Small for Gestational Age: Differential Diagnosis, Molecular Genetic Evaluation, and Implications. Endocr Rev. Dec 1 2018;39(6):851–894. doi:10.1210/er.2018-00083

15. McCowan L, Horgan RP. Risk factors for small for gestational age infants. Best Pract Res Clin Obstet Gynaecol. Dec 2009;23(6):779–93. doi:10.1016/j.bpobgyn.2009.06.003

16. Wollmann HA. Intrauterine growth restriction: definition and etiology. Horm Res. 1998;49 Suppl 2:1–6. doi:10.1159/000053079

17. Mone F, Mellis R, Gabriel H, et al. Should we offer prenatal exome sequencing for intrauterine growth restriction or short long bones? A systematic review and meta-analysis. Am J Obstet Gynecol. Oct 7 2022;doi:10.1016/j.ajog.2022.09.045

18. Meler E, Sisterna S, Borrell A. Genetic syndromes associated with isolated fetal growth restriction. Prenat Diagn. Mar 2020;40(4):432–446. doi:10.1002/pd.5635

19. Peng R, Yang J, Xie HN, Lin MF, Zheng J. Chromosomal and subchromosomal anomalies associated to small for gestational age fetuses with no additional structural anomalies. Prenat Diagn. Dec 2017;37(12):1219–1224. doi:10.1002/pd.5169

20. Borrell A, Grande M, Meler E, et al. Genomic Microarray in Fetuses with Early Growth Restriction: A Multicenter Study. Fetal Diagn Ther. 2017;42(3):174–180. doi:10.1159/000452217

21. Motelow JE, Lippa NC, Hostyk J, et al. Risk Variants in the Exomes of Children With Critical Illness. JAMA Netw Open. Oct 3 2022;5(10):e2239122. doi:10.1001/jamanetworkopen.2022.39122

22. Stanley KE, Giordano J, Thorsten V, et al. Causal Genetic Variants in Stillbirth. N Engl J Med. Sep 17 2020;383(12):1107–1116. doi:10.1056/NEJMoa1908753

23. Petrovski S, Aggarwal V, Giordano JL, et al. Whole-exome sequencing in the evaluation of fetal structural anomalies: a prospective cohort study. Lancet. Feb 23 2019;393(10173):758–767. doi:10.1016/S0140-6736(18)32042-7

24. Lord J, McMullan DJ, Eberhardt RY, et al. Prenatal exome sequencing analysis in fetal structural anomalies detected by ultrasonography (PAGE): a cohort study. Lancet. Feb 23 2019;393(10173):747–757. doi:10.1016/S0140-6736(18)31940-8

25. Bertoli-Avella AM, Beetz C, Ameziane N, et al. Successful application of genome sequencing in a diagnostic setting: 1007 index cases from a clinically heterogeneous cohort. Eur J Hum Genet. Jan 2021;29(1):141–153. doi:10.1038/s41431-020-00713-9

26. Hays T, Wapner RJ. Genetic testing for unexplained perinatal disorders. Curr Opin Pediatr. Apr 1 2021;33(2):195–202. doi:10.1097/MOP.0000000000000999

27. Spitzer AR, Ellsbury DL, Handler D, Clark RH. The Pediatrix BabySteps Data Warehouse and the Pediatrix QualitySteps improvement project system--tools for “meaningful use” in continuous quality improvement. Clin Perinatol. Mar 2010;37(1):49–70. doi:10.1016/j.clp.2010.01.016

28. Grantz KL, Grewal J, Kim S, et al. Unified standard for fetal growth: the Eunice Kennedy Shriver National Institute of Child Health and Human Development Fetal Growth Studies. Am J Obstet Gynecol. Apr 2022;226(4):576–587.e2. doi:10.1016/j.ajog.2021.12.006

29. Tolia VN, Clark RH. The Denominator Matters! Lessons from Large Database Research in Neonatology. Children (Basel). Nov 7 2020;7(11):216–216. doi:10.3390/children7110216

30. Papile LA, Burstein J, Burstein R, Koffler H. Incidence and evolution of subependymal and intraventricular hemorrhage: a study of infants with birth weights less than 1,500 gm. J Pediatr. Apr 1978;92(4):529–34. doi:10.1016/s0022-3476(78)80282-0

31. Stevenson DK, Verter J, Fanaroff AA, et al. Sex differences in outcomes of very low birthweight infants: the newborn male disadvantage. Arch Dis Child Fetal Neonatal Ed. Nov 2000;83(3):F182–5. doi:10.1136/fn.83.3.f182

32. Peacock JL, Marston L, Marlow N, Calvert SA, Greenough A. Neonatal and infant outcome in boys and girls born very prematurely. Pediatr Res. Mar 2012;71(3):305–10. doi:10.1038/pr.2011.50

33. *R: A language and environment for statistical computing*. R Foundation for Statistical Computing; 2022. https://www.R-project.org/

34. Schlüter DK, Griffiths R, Adam A, et al. Impact of cystic fibrosis on birthweight: a population based study of children in Denmark and Wales. Thorax. 2019;74(5):447–454. doi:10.1136/thoraxjnl-2018-211706

35. Giorgione V, Briffa C, Di Fabrizio C, Bhate R, Khalil A. Perinatal Outcomes of Small for Gestational Age in Twin Pregnancies: Twin vs. Singleton Charts. J Clin Med. Feb 8 2021;10(4) doi:10.3390/jcm10040643

36. D’Antonio F, Khalil A, Dias T, Thilaganathan B. Weight discordance and perinatal mortality in twins: analysis of the Southwest Thames Obstetric Research Collaborative (STORK) multiple pregnancy cohort. Ultrasound Obstet Gynecol. Jun 2013;41(6):643–8. doi:10.1002/uog.12412

37. Townsend R, Khalil A. Fetal growth restriction in twins. Best Pract Res Clin Obstet Gynaecol. May 2018;49:79–88. doi:10.1016/j.bpobgyn.2018.02.004

38. Kim F, Bateman DA, Goldshtrom N, Sheen J-J, Garey D. Intracranial ultrasound abnormalities and mortality in preterm infants with and without fetal growth restriction stratified by fetal Doppler study results. Journal of Perinatology. 2023/01/30 2023;doi:10.1038/s41372-023-01621-8

39. Procianoy RS, Garcia-Prats JA, Adams JM, Silvers A, Rudolph AJ. Hyaline membrane disease and intraventricular haemorrhage in small for gestational age infants. Arch Dis Child. Jul 1980;55(7):502–5. doi:10.1136/adc.55.7.502

40. Gilbert WM, Danielsen B. Pregnancy outcomes associated with intrauterine growth restriction. American Journal of Obstetrics and Gynecology. 2003/06/01/ 2003;188(6):1596–1601. doi:https://doi.org/10.1067/mob.2003.384

41. Sacchi C, Marino C, Nosarti C, Vieno A, Visentin S, Simonelli A. Association of Intrauterine Growth Restriction and Small for Gestational Age Status With Childhood Cognitive Outcomes: A Systematic Review and Meta-analysis. JAMA Pediatr. Aug 1 2020;174(8):772–781. doi:10.1001/jamapediatrics.2020.1097

42. Roberts D, Brown J, Medley N, Dalziel SR. Antenatal corticosteroids for accelerating fetal lung maturation for women at risk of preterm birth. Cochrane Database Syst Rev. Mar 21 2017;3(3):Cd004454. doi:10.1002/14651858.CD004454.pub3

43. Blankenship SA, Brown KE, Simon LE, Stout MJ, Tuuli MG. Antenatal corticosteroids in preterm small-for-gestational age infants: a systematic review and meta-analysis. Am J Obstet Gynecol MFM. Nov 2020;2(4):100215. doi:10.1016/j.ajogmf.2020.100215

44. Schmidt B, Davis P, Moddemann D, et al. Long-term effects of indomethacin prophylaxis in extremely-low-birth-weight infants. N Engl J Med. Jun 28 2001;344(26):1966–72. doi:10.1056/nejm200106283442602

45. Mirza H, Laptook AR, Oh W, et al. Effects of indomethacin prophylaxis timing on intraventricular haemorrhage and patent ductus arteriosus in extremely low birth weight infants. Arch Dis Child Fetal Neonatal Ed. Sep 2016;101(5):F418–22. doi:10.1136/archdischild-2015-309112

46. Foglia EE, Roberts RS, Stoller JZ, Davis PG, Haslam R, Schmidt B. Effect of Prophylactic Indomethacin in Extremely Low Birth Weight Infants Based on the Predicted Risk of Severe Intraventricular Hemorrhage. Neonatology. 2018;113(2):183–186. doi:10.1159/000485172

47. Fetal Growth Restriction: ACOG Practice Bulletin, Number 227. Obstet Gynecol. Feb 1 2021;137(2):e16–e28. doi:10.1097/aog.0000000000004251

48. Martins JG, Biggio JR, Abuhamad A. Society for Maternal-Fetal Medicine Consult Series #52: Diagnosis and management of fetal growth restriction: (Replaces Clinical Guideline Number 3, April 2012). Am J Obstet Gynecol. Oct 2020;223(4):B2–b17. doi:10.1016/j.ajog.2020.05.010

49. Dauber A. Genetic Testing for the Child With Short Stature-Has the Time Come To Change Our Diagnostic Paradigm? J Clin Endocrinol Metab. Jul 1 2019;104(7):2766–2769. doi:10.1210/jc.2019-00019

50. Rapaport R, Wit JM, Savage MO. Growth failure: ‘idiopathic’ only after a detailed diagnostic evaluation. Endocr Connect. Mar 2021;10(3):R125–r138. doi:10.1530/ec-20-0585

51. Clark MM, Stark Z, Farnaes L, et al. Meta-analysis of the diagnostic and clinical utility of genome and exome sequencing and chromosomal microarray in children with suspected genetic diseases. NPJ Genom Med. 2018;3(1):16. doi:10.1038/s41525-018-0053-8

52. Wojcik MH, Schwartz TS, Yamin I, et al. Genetic disorders and mortality in infancy and early childhood: delayed diagnoses and missed opportunities. Genet Med. Nov 2018;20(11):1396–1404. doi:10.1038/gim.2018.17

